# Enhanced Diabetes Prediction Using Novel Additive-Multiplicative Neural Networks: A Comprehensive Machine Learning Analysis of the PIMA Indians Dataset

**DOI:** 10.1101/2025.09.20.25336250

**Authors:** Şeyda Demirel Tatli, Kursat Aytekin, Melih Agraz

## Abstract

**Background:** Early diabetes detection remains challenging, requiring robust machine learning approaches that balance accuracy with clinical interpretability for effective diagnostic support.

**Methods:** We are proposing a novel Additive and Multiplicative Neurons Network (AMNN) that combines both additive and multiplicative computational pathways to capture complex nonlinear relationships in diabetes prediction. Using the PIMA Indians Diabetes dataset (n=768), we compared AMNN against nine established algorithms including XGBoost, KAN, and traditional neural networks. Data preprocessing included SMOTE oversampling for class imbalance, and model interpretability was enhanced through SHAP and LIME explainable AI techniques.

**Results:** The AMNN model outperformed all baseline approaches, achieving 75.76% accuracy, a 76.18% F1-score, and an AUC-ROC of 0.8206. Across both traditional feature selection techniques and explainable AI analyses, glucose levels, BMI, age, and pregnancy count consistently emerged as the most influential predictors.

**Conclusions:** The AMNN framework demonstrates strong potential for diabetes prediction by balancing accuracy with clinical interpretability. The key predictors it highlights align closely with established medical knowledge, reinforcing confidence in its outputs and suitability for use in clinical decision-making workflows. This hybrid neural network approach represents a promising step toward transparent, AI-assisted diagnostic tools that can support healthcare professionals in practice.

## 1. INTRODUCTION

Diabetes develops when the body either cannot use insulin effectively or fails to produce enough of it to regulate blood glucose levels. This condition increases the risk of serious complications such as heart and kidney disease, vascular and nerve damage, and even blindness [1]. As technology continues to advance, artificial intelligence (AI) and machine learning (ML) have become powerful tools in transforming medical research and practice. They are widely applied in areas such as genetics [43], drug discovery [44], and disease diagnosis [21]. Machine learning techniques offer significant advantages: they speed up diagnostic processes, reduce patient waiting times in healthcare settings, and streamline healthcare operations by automating tasks that would otherwise require human labor. By rapidly analyzing large and complex datasets, ML algorithms deliver accurate results at a lower cost. For these reasons, machine learning has become a preferred approach among researchers for disease diagnosis [2].

Recent advancements also emphasize the growing importance of model interpretability and biomarker optimization in medical studies. For instance, the Extended lasso-type MARS (LMARS) model has been used to describe biological networks, offering a flexible framework for uncovering hidden structures in complex biomedical data [47]. Similarly, ChatGPT-Enhanced ROC Analysis (CERA) provides a novel web-based tool for determining optimal cutoff points in biomarker analysis, thereby supporting more precise diagnostic decision-making [48]. Both studies complement the growing body of work that integrates advanced statistical modeling and AI-enhanced tools into healthcare, highlighting how novel methods can strengthen disease diagnosis and predictive accuracy in conditions such as diabetes.

In this study, publicly available [3] PIMA dataset was used for machine learning algorithms in which the dataset was originally obtained from the National Institute of Diabetes and Digestive and Kidney Diseases. The study is designed to predict whether an individual has diabetes based on a range of diagnostic indicators, while also highlighting the factors that contribute to diabetes. Notably, all participants in the dataset are Pima Indian women aged 21 or above [3]. There is extensive research in the literature related to the PIMA Diabetes dataset. Kayaer and Yildirim [4] examined the General Regression Neural Network (GRNN) structure on this dataset and found that it achieved similar classification accuracy to more complex neural networks, such as Multi-Layer Perceptron (MLP) and RBF, obtaining an accuracy rate of 80.21% for GRNN and 81% for ARTMAP-IC.4. Karatsiolis and Schizas [5] aimed to improve these accuracy rates by using a modified Support Vector Machine (SVM) strategy, achieving an accuracy of 82.2% for diabetes classification on the Pima Indians Diabetes dataset. Yangın [6] compared decision trees, Random Forest (RF), Gradient Boosting, and XGBoost (eXtreme Gradient Boosting) machine learning algorithms on two different datasets, including the Pima Diabetes dataset, and identified the highest classification accuracy with 82.35%. Ganesh and Sripriya [7] examined different classification methods on the Pima Indians Diabetes dataset, evaluating the advantages and disadvantages of techniques used for diabetes prediction and demonstrating that preprocessing improves classification accuracy. Lakhwani et al. [8] developed an artificial neural network (ANN)-based automated diabetes diagnosis system using the same dataset and used cumulative gain charts to measure model quality. Patra and Khuntia [9] improved the k-nearest neighbors (KNN) classifier using a new standard deviation-based distance calculation method on the Pima Indians Diabetes dataset, achieving an accuracy of 83.2%. Mousa et al. [10] compared Long Short-Term Memory (LSTM), RF, and Convolutional Neural Network (CNN) models for detecting diabetes using the same dataset, showing that LSTM achieved the highest accuracy (85%) and was particularly effective in capturing temporal patterns. Chang et al. [11] developed an e-diagnosis framework using the dataset evaluating interpretable ML models such as Naïve Bayes, RF, and J48. Their analysis showed RF attained the best accuracy (79.57%), though Naïve Bayes performed well with fewer features and J48 maintained strong sensitivity. In addition, Farsana and Poulo [12] analyzed the PIMA dataset by comparing hybrid CNN models with traditional machine learning approaches to aid in the early diagnosis and prevention of diabetes. Their hybrid CNN model attained an accuracy of 73%.

The main aim of this study is to improve diabetes prediction by applying different machine learning approaches and determining the most influential variables in classification. To accomplish this, multiple models were trained and assessed, with the optimal algorithm selected according to critical performance metrics. Furthermore, feature importance was analyzed using both feature selection techniques and explainable artificial intelligence (XAI) methods. The primary innovation of this study is the implementation of the Additive and Multiplicative Neurons Network (AMNN), which outperformed all other models. Additionally, the study introduces the KAN model as an alternative approach to exploring flexible neural architectures.

The subsequent parts of this research are arranged in the following manner: The second section introduces the Pima Indians Diabetes dataset and describes the preprocessing steps applied to the dataset. The third section discusses model selection criteria and presents the selected models. The fourth section compares the methods used in the study and reports the findings. Finally, the last section presents the conclusions and recommendations based on the obtained results.

## 2. METHODS

### 2.1. Dataset

In the PIMA dataset, there are 768 instances described by nine variables. Among the 768 individuals in the dataset, 268 have diabetes, while 500 do not. The independent variables for diabetic individuals are described in Table 1.

**Table 1.** Variables in the dataset.

| Variable | Unit | Description |
| --- | --- | --- |
| Pregnancies | Count | Number of pregnancies |
| Glucose | mg/dL | 2-hour plasma glucose concentration |
| Blood Pressure | mm Hg | Diastolic blood pressure |
| Skin Thickness | Mm | Triceps skin fold thickness |
| Insulin | $\mu\text{U/ml}$ | 2-hour serum insulin level |
| BMI | $\text{kg}/(\text{m}^2)$ | Body mass index (BMI) |
| Diabetes Pedigree Function | Numeric | Diabetes pedigree function |
| Age | Years | Age of the individual |
| Outcome | Categorical | Class label (0: non-diabetic, 1: diabetic) |

To assess the suitability of the dataset, we draw a t-SNE plot, as shown in Figure 1.

**Figure 1.**
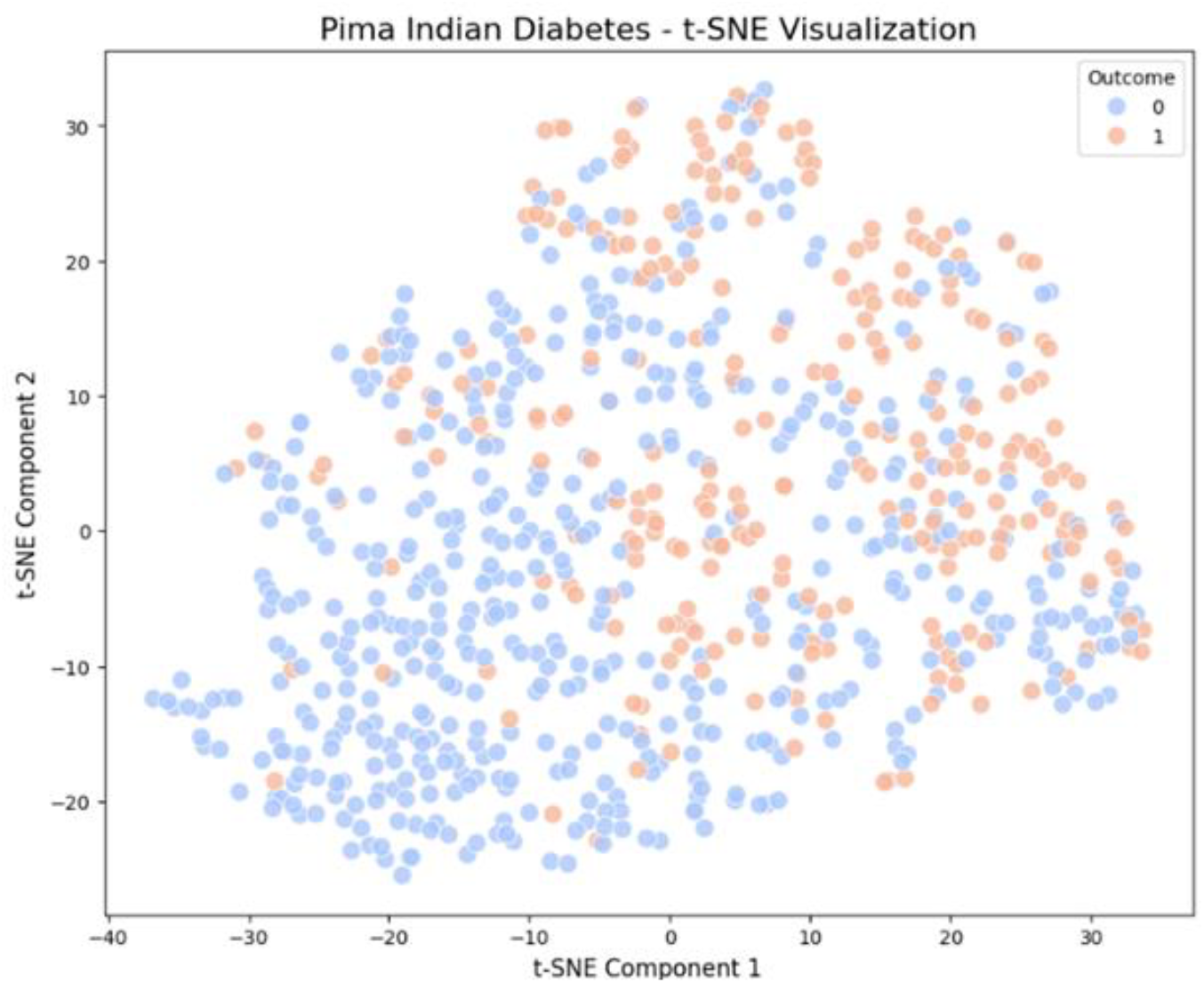
Two-dimensional t-SNE visualization of the PIMA Indians Diabetes dataset showing the distribution and separability of diabetic (orange) and non-diabetic (blue) cases.

The plot demonstrates partial clustering with some overlap between classes, indicating the dataset’s complexity and the need for sophisticated classification approaches.

As seen in Figure 1, the dataset exhibits certain clustering patterns, with some data points forming distinct clusters while others appear more scattered. The colors in the visualization indicate that data points labeled as Outcome = 0/no-diabetes (blue) and Outcome = 1/diabetes (orange) are somewhat distinguishable but not entirely separable.

### 2.2. Data Preprocessing

The dataset was preprocessed using Python (version 3.10.12). The summary statistics of the dataset are reported in Table 2.

**Table 2.** Descriptive statistics of the data.

| Variable | Mean | Std | Min | 25% | 50% | 75% | Max |
| --- | --- | --- | --- | --- | --- | --- | --- |
| Pregnancies | 3,85 | 3,37 | 0,00 | 1,00 | 3,00 | 6,00 | 17,00 |
| Glucose | 120,89 | 31,97 | 0,00 | 99,00 | 117,00 | 140,25 | 199,00 |
| Blood Pressure | 69,11 | 19,36 | 0,00 | 62,00 | 72,00 | 80,00 | 122,00 |
| Skin Thickness | 20,54 | 15,95 | 0,00 | 0,00 | 23,00 | 32,00 | 99,00 |
| Insulin | 79,80 | 115,24 | 0,00 | 0,00 | 30,50 | 127,25 | 846,00 |
| BMI | 31,99 | 7,88 | 0,00 | 27,30 | 32,00 | 36,60 | 67,10 |
| Pedigree | 0,47 | 0,33 | 0,08 | 0,24 | 0,37 | 0,63 | 2,42 |
| Age | 33,24 | 11,76 | 21,00 | 24,00 | 29,00 | 41,00 | 81,00 |

Additionally, the following data preprocessing methods were applied throughout the entire analysis Missing Data: Although there are no explicitly missing values in the dataset, certain variables contain an excessive number of zero values, which are considered missing. These zero values were treated as missing observations and imputed using the median.

Data Scaling: An analysis of the dataset revealed that variables have different magnitudes. For instance, the Age variable ranges from 21 to 81, whereas the Diabetes Pedigree Function variable ranges from 0.07 to 2.42. To eliminate these differences, Min-Max scaling were applied.

Imbalanced Data: Refers to datasets in which the distribution of classes is markedly unequal, a common challenge in classification problems [13]. In the Pima dataset, there is a pronounced imbalance in class representation. In the Pima dataset, there is a noticeable difference between diabetic (34.9%) and non-diabetic (65.1%) individuals. This class imbalance can hinder the accurate prediction of the minority class (diabetic individuals). The imbalance issue was addressed by applying the oversampling (SMOTE) approach during the stages of model and feature selection processes [14].

### 2.3. Feature Engineering

#### 2.3.1. Feature Selection Methods

Correlation: When selecting features, it is essential to examine the relationships between variables. Independent variables should be highly correlated with the dependent variable; however, high correlations among independent variables can cause multicollinearity, which is undesirable. One of the feature selection techniques, CFS (Correlation Feature Selection), determines a subset of features using a correlation matrix. When analyzing the correlation matrix, it is observed that the variables with a correlation value of 0.20^+^with the outcome variable are [‘Glucose’, ‘BMI’, ‘Pregnancies’, ‘Age’]. We plot the correlation matrix, as seen in Figure 2 below.

**Figure 2.**
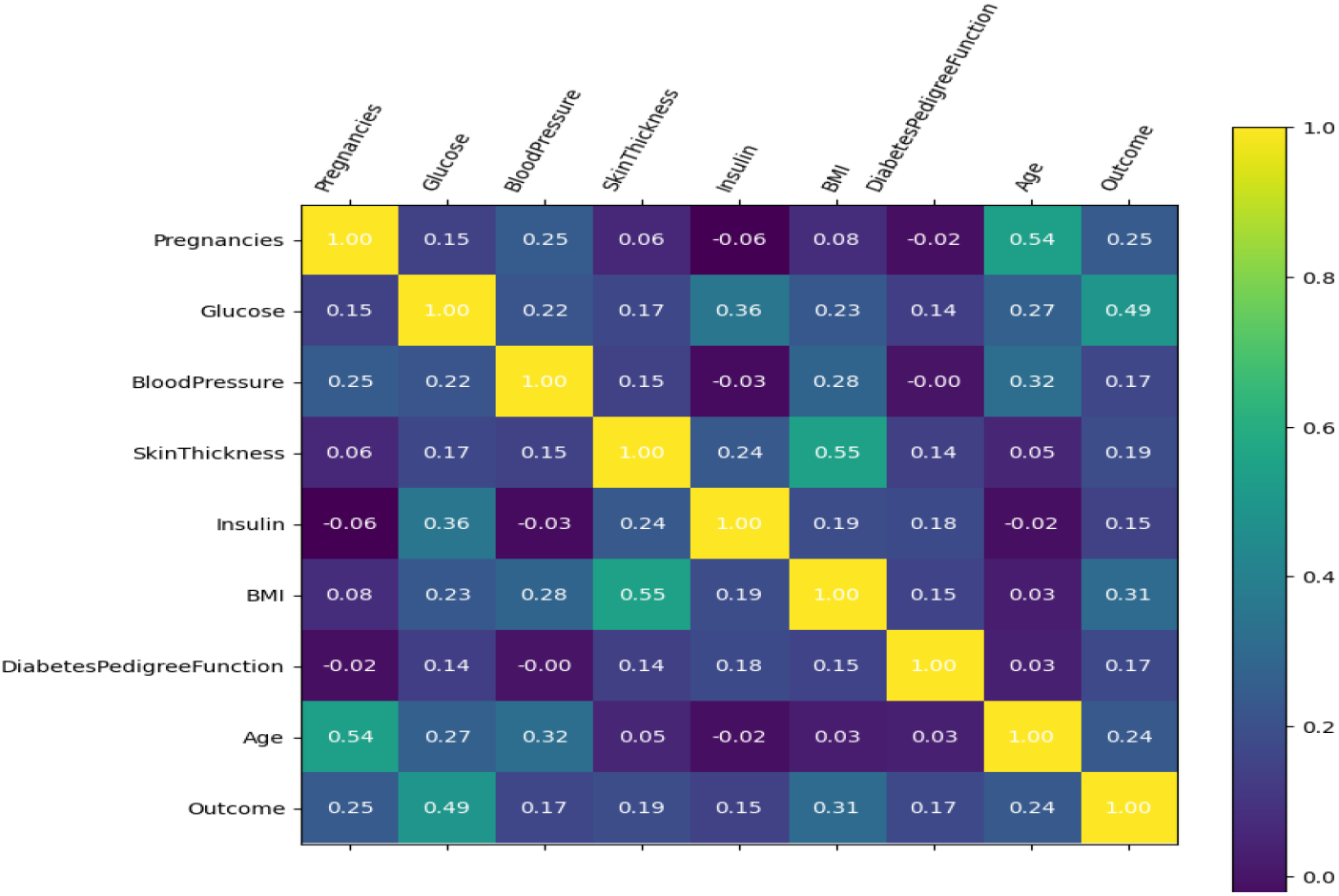
Pearson correlation matrix heatmap for all variables in the PIMA dataset.

Boruta: Another feature selection method used is Boruta [15]. This method analyzes the relationships between dependent and independent variables to determine which variables are important for the predictive model. Based on the Random Forest (RF) algorithm, Boruta compares the relevance of each variable with artificially generated ‘shadow’ variables. Through this comparison, it identifies the most significant real features [16] Selected important features: [‘Pregnancies’, ‘Glucose’, ‘BMI’, ‘DiabetesPedigreeFunction’, ‘Age’].

Maximum Relevance - Minimum Redundancy (MRMR): which aims to choose features that are strongly associated with the target variable while minimizing redundancy among them [17]. This method selects the most relevant features to the output while ensuring minimal similarity between selected features. The formula for this method is given in Equation 1 [18].

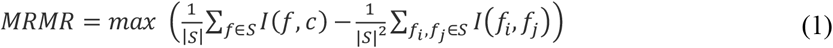

Selected important features: [‘Glucose’, ‘BMI’, ‘Pregnancies’, ‘DiabetesPedigreeFunction’, ‘Age’] Recursive Feature Elimination (RFE): This iterative process eliminates the least important features to select the best-performing set of features. It begins by training step with all available features. Then, in each iteration, it identifies and eliminates the feature with the least significance, retraining the model with the other features. In summary, at each iteration, the weakest feature is removed, and the model is retrained until the predefined number of features is obtained [19]. Feature ranking using RFE: [1, 1, 2, 3, 4, 1, 1, 1]. Selected important features: [‘Pregnancies’, ‘Glucose’, ‘BMI’, ‘DiabetesPedigreeFunction’, ‘Age’]

Random Forest (RF): RF itself is another feature selection strategy since it is an ensemble method based on multiple decision trees. RF, the importance scores of variables are calculated, and high-scoring features are retained in the model. It assesses each feature’s significance by examining its influence on the splits within decision trees, using measures like Gini index or entropy gain [20,25]. The feature importance scores from the RF method are provided in the appendix, but the five most important features are [‘Glucose’, ‘BMI’, ‘Age’, ‘DiabetesPedigreeFunction’, and ‘Pregnancies’].

A comparison of the results obtained from these five feature selection techniques is summarized in Table 3.

**Table 3.** Features selected by different methods.

| Methods | Features Selected |  |  |  |  |
| --- | --- | --- | --- | --- | --- |
| Correlation | Glucose | BMI | Age | Pregnancies |  |
| Boruta | Glucose | BMI | Pregnancies | PedigreeFunction | Age |
| MRMR | Glucose | BMI | Pregnancies | PedigreeFunction | Age |
| RFE | Glucose | BMI | Pregnancies | PedigreeFunction | Age |
| Random Forest | Glucose | BMI | Age | PedigreeFunction | Pregnancies |

Using the majority voting method [22] it is observed that ‘Glucose’, ‘BMI’, ‘Age’, and ‘Pregnancies’ were consistently selected across all methods, highlighting their significance for the model. In methods where feature ranking is important (Correlation, MRMR, and RF), “Glucose” and “BMI” were ranked among the top two features, while the rankings of other features varied by method. However, since ranking is not crucial for Boruta and RFE, features were selected based on their presence rather than their order. Based on these observations, “Glucose” and “BMI” are identified as the most critical features for diabetes diagnosis, followed by “Pregnancies”, “DiabetesPedigreeFunction”, and “Age”.

#### 2.3.2. Explainable Machine Learning (XAI) Approaches

Explainable machine learning (XAI) methods aim to make model decisions understandable and interpretable. After training a model on the Pima dataset, widely used XAI methods such as SHAP (SHapley Additive ExPlanations) or LIME (Local Interpretable Model-Agnostic Explanations) [23]. The SHAP summary plot is provided in Figure 3.

**Figure 3.**
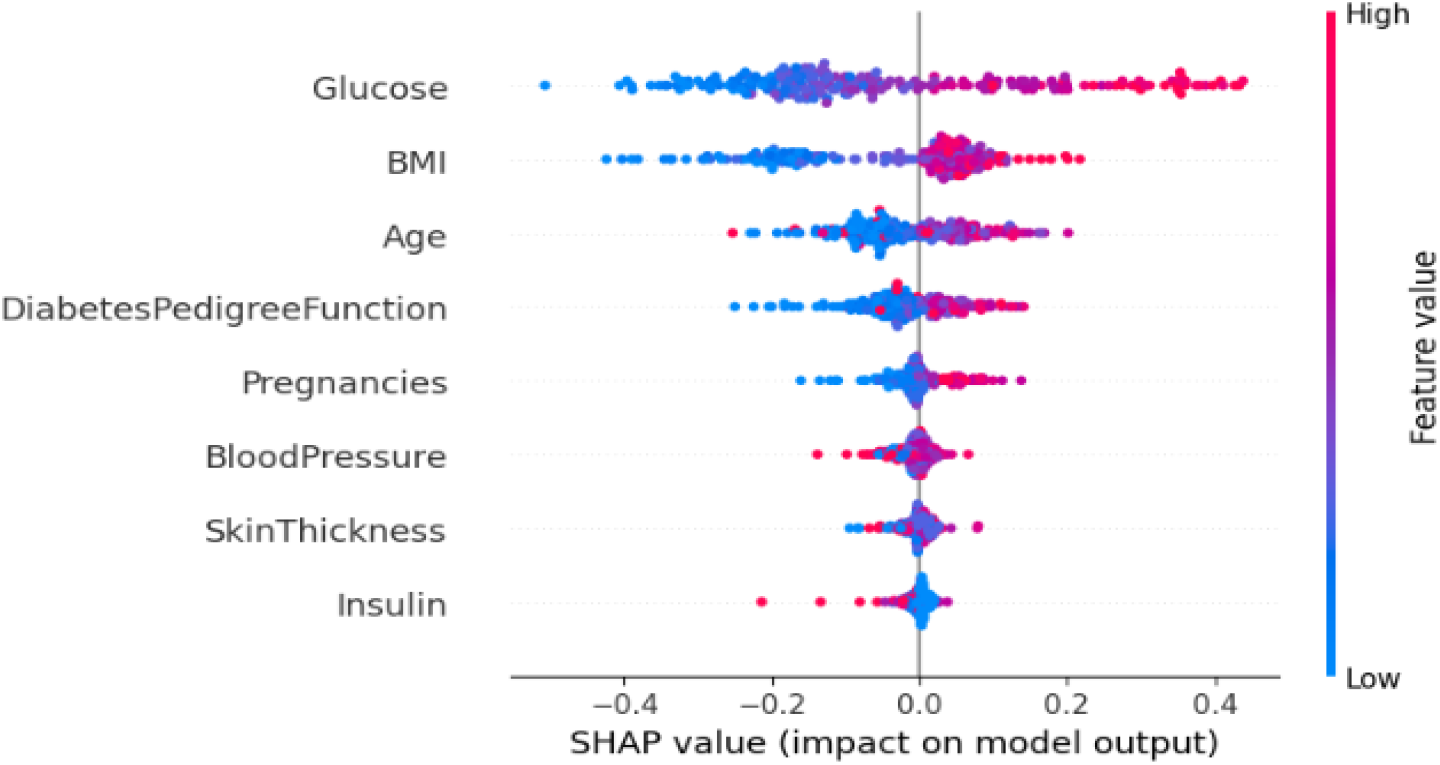
SHAP (SHapley Additive exPlanations) summary plot displaying feature importance and impact direction for diabetes prediction. Each point represents a patient, with red indicating high feature values and blue indicating low values. Glucose emerges as the most influential predictor, followed by BMI, Age, and Pregnancies. Horizontal spread indicates the magnitude of each feature’s impact on model predictions.

When analyzing the SHAP plot, it is observed that “**Glucose**” is the most influential feature for diabetes diagnosis. Higher glucose levels (represented by red points) substantially raise the likelihood of a diabetes diagnosis, whereas lower glucose levels (blue points) reduce this probability. After glucose, the next most influential feature is “**BMI**”, which also plays a significant role in predicting diabetes risk. As BMI increases, the model assigns a higher likelihood of diabetes diagnosis, highlighting the importance of weight management in diabetes prediction.

#### 2.3.3. Toy LIME Example on Patient 163

LIME is a model-agnostic technique that provides interpretability by approximating complex model predictions locally with simpler, interpretable models, thus offering insights into the factors influencing individual predictions. LIME visualization is presented on the patient number 163 in Figure 4.

**Figure 4.**
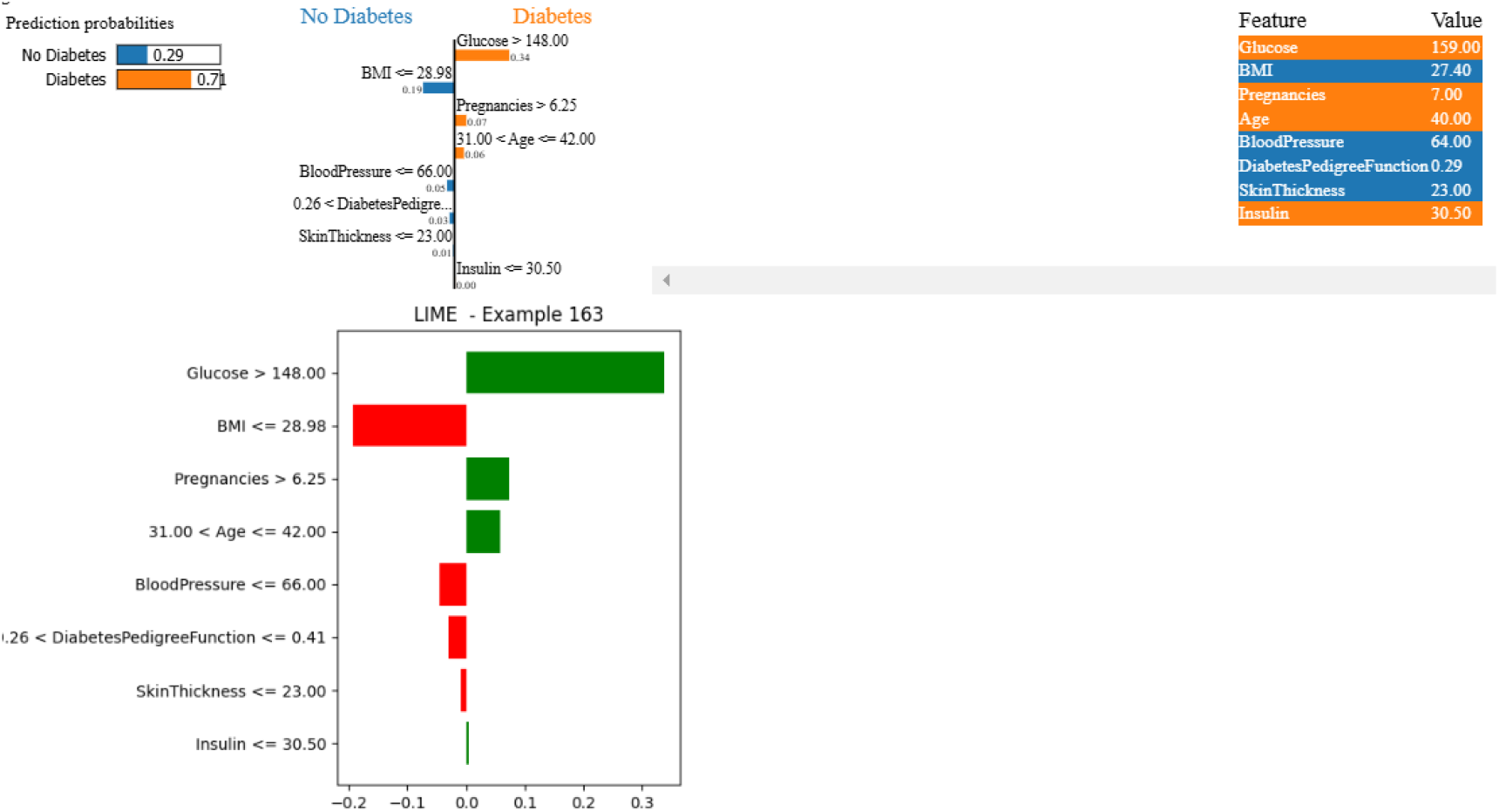
LIME (Local Interpretable Model-agnostic Explanations) analysis for Patient 163, demonstrating individual prediction explanations. The visualization shows feature contributions to diabetes risk prediction, with BMI <28.98 contributing to 19% risk reduction, while Glucose >148 increases risk by 34%. This patient-specific analysis illustrates how different clinical parameters influence individual diagnostic predictions.

According to the LIME results on patient number 163, BMI, Glucose level, Age, and Number of Pregnancies significantly influence the risk of diabetes. A BMI below 28.98 contributes to a 19% reduction in risk, while a BMI above this threshold is related with a higher risk of diabetes. Glucose level is the most influential factor, with a 34% effect; high glucose levels (above 148) substantially increase the risk of diabetes. The age range of 31-42 years contributes to a 6% increase in risk, and the number of pregnancies above 6.25 has a 7% effect on diabetes risk. DiabetesPedigreeFunction and other factors such as Skin Thickness and Insulin level have less significant contributions to the overall risk. These findings suggest that while glucose level and BMI are the strongest predictors, age and pregnancy history also play a role, though to a lesser extent.

### 2.4. Model Selection

Model selection in data mining is critical for identifying the best model for a given task. This process often involves evaluation criteria that measure accuracy, generalizability, and efficiency. Commonly used metrics include accuracy, the F1-score, cross-validation performance, and the area under the ROC curve (AUC-ROC). It is also important to consider model complexity and parameter tuning to avoid overfitting or underfitting. In addition, the efficiency and speed of the model play a significant role in the selection process.

To evaluate how well different models perform, a confusion matrix provides a visual summary of predictions versus actual outcomes. Table 4 illustrates the confusion matrix for a binary classification scenario, with the detailed results provided below

**Table 4.** Confusion matrix for the binary classification problem.

| CONFUSION MATRIX |  | ACTUAL VALUE |  |
| --- | --- | --- | --- |
|  |  | Positive: Diabetes | Negative: No Diabetes |
| PREDICTED VALUE | Positive: Diabetes | TP | FP |
|  | Negative: No Diabetes | FN | TN |

- True Positive (TP): Cases that are actually positive and are correctly predicted as positive by the model.
- False Positive (FP): Cases that are actually negative but are incorrectly predicted as positive by the model.
- True Negative (TN): Cases that are actually negative and are correctly predicted as negative.
- False Negative (FN): ases that are actually positive but are incorrectly predicted as negative.

Based on the matrix presented in Table 4, the corresponding performance metrics are calculated. The performance measures provided sequentially are accuracy, precision, recall (sensitivity), F1-score, and ROC curve.

#### Accuracy

Defined as the fraction of instances that are correctly classified relative to the total number of classified instances. It is commonly used as a measure of prediction performance in classification problems. However, using accuracy alone can be misleading, so employing multiple performance metrics provides a more comprehensive evaluation. 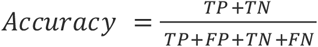

#### Precision

Precision measures the proportion of correctly predicted positive instances relative to all instances classified as positive. 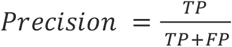

#### Recall (Sensitivity)

Calculates the model’s success to correctly identify real positive labels in the test set. It is defined as the ratio of true positives to all actual positives. 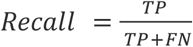

#### F1-Score

Since working with two different metrics can sometimes be problematic, researchers may prefer a single number to measure performance. F1-score is calculated as a combination of recall and precision. 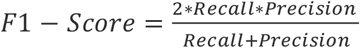

#### ROC Curve

The Receiver Operating Characteristic (ROC) curve is a probability curve for different classes. The x-axis presents the false positive ratio, while the y-axis presents the true positive predictions. This curve illustrates how well the classifier performs in making predictions. The area under the curve (AUC) ranges from 0 to 1, with higher values indicating better discriminatory ability.In binary classification, the ROC curve provides a visual summary of the trade-off between correctly identifying positive instances and the risk of misclassifying negatives as positives. Overall, ROC curves are valuable for comparing the effectiveness of different classifiers [24].

### 2.5. Machine Learning Methods

Classification problems aim to determine the class of a new observation based on the analysis of a given dataset. Various methods can be used for solving classification problems. In this study, machine learning methods frequently used in the literature for diabetes diagnosis are considered.

Logistic Regresyon (LR): A statistical/machine learning model used in binary classification problems where the dependent variable is discrete. It is widely applied in computer science, various applied sciences, and real-world problems. Logistic regression explains the relation between a binary dependent variable and a set of independent variables using a logistic function (logit function) [26]. The probability of an event occurring is given by Equation 2. The logit function is calculated as Equation 3 [27].

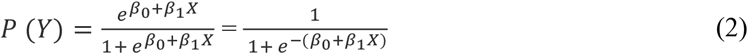

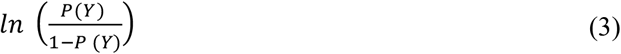

Multi-Layer Perceptron (MLP): An MLP is a fundamental type of artificial neural network with a feedforward architecture, typically composed of at least one hidden layer. The structure features an input layer, hidden layers, and an output layer, all linked through full connectivity. MLPs utilize nonlinear activation functions such as ReLU, Sigmoid, and Tanh to catch complex patterns and relationships. They are trained through the backpropagation algorithm, and their weights are updated using optimization techniques based on gradient descent. MLPs are extensively used for classification, regression, and time series prediction tasks, particularly when addressing nonlinear problems [28].

Extreme Gradient Boosting (XGBoost): XGBoost is a high-performance, optimized gradient boosting algorithm. It builds upon the gradient boosting framework originally proposed by Friedman [29] and and includes various enhancements that make it especially effective for large datasets and high-dimensional feature spaces. XGBoost has shown superior performance across a variety of applications [30].

Extra-Trees (ET): Extra-Trees is an ensemble learning method developed by Geurts [31] as an extension of the Random Forest algorithm proposed by Breiman [20]. It constructs multiple decision trees while increasing randomness to prevent overfitting. Unlike Random Forest, Extra-Trees selects split points randomly, leading to faster tree construction. This approach reduces computational cost while maintaining generalization performance. Extra-Trees is used in classification, regression, and feature importance evaluation, and it performs well on noisy datasets [31].

K-Nearest Neighbors (KNN): KNN is a non-parametric approach introduced by Fix and Hodges [32] and commonly employed for both classification and regression tasks. In classification, it determines a label by looking at the K nearest neighbors and choosing the class that appears most frequently among them. Since KNN searches for the closest neighbors for each data point, the computational complexity increases as the dataset grows. As KNN is distance-based, normalizing the training data can significantly improve its accuracy.

Gaussian Naive Bayes (GNB): GNB is a type of Naive Bayes classifier that assumes a Gaussian (Normal) distribution for continuous data. The algorithm relies on Bayes’ Theorem to compute conditional probabilities and make class predictions. GNB operates under the assumption that features are independent of one another, which simplifies the model and allows for fast and computationally efficient processing.. Since GNB considers features to be normally distributed, it estimates class probabilities using the mean and variance of each feature. GNB is particularly effective for high-dimensional datasets and small sample sizes. It is widely applied in text classification, medical diagnosis, fraud detection [33].

Kolmogorov-Arnold Networks (KAN): KANs are a type of neural network that employ a fundamentally different learning approach compared to traditional multilayer perceptrons (MLPs). Unlike MLPs, which use fixed activation functions at each neuron, KANs assign adaptable activation functions to the connections between nodes. This modification, though subtle in appearance, can significantly improve both the network’s performance and its interpretability. In KANs, each connection weight is replaced by a univariate function-often implemented as a spline-thereby removing the reliance on linear weights. The nodes themselves simply aggregate incoming signals without performing any nonlinear transformations. [34].

#### 2.5.1 Artificial Neural Network Approach with Additive and Multiplicative Neurons (AMNN)

Deep Learning-Based Alternative Model: Neural Network Approach with Additive and Multiplicative Neurons (AMNN): Traditional neural networks generally use additive operations, with each neuron computing the weighted sum of incoming signals and applying an activation function. However, this structure may fall short in modeling complex nonlinear relationships in certain applications. To address this limitation, more flexible models have been developed that integrate both additive and multiplicative neurons. AMNN offer an enhanced ability to learn richer representations by employing both summation (∑) and multiplication (∏) operations on the inputs. Specifically, multiplicative neurons are beneficial for modeling interactions between inputs. This approach helps overcome the limited linear combination capability of traditional neural networks, enabling the model to capture more intricate relationships. Models utilizing both additive and multiplicative neurons have been developed as an alternative to conventional ANNs, aiming to achieve more effective results in specific problems by allowing the network to learn more complex dependencies between input features [45, 46].

As illustrated in the model architecture diagram in Figure 5, the network comprises two parallel paths originating from the input layer. The first path includes a conventional neuron that performs a weighted summation (i.e., linear combination), while the second path consists of a computational unit that multiplies all input features together. These two pathways are subsequently merged through summation to generate a unified representation. A hidden layer with ReLU activation follows this combined output, and finally, a sigmoid-activated output neuron is employed for binary classification.

**Figure 5.**
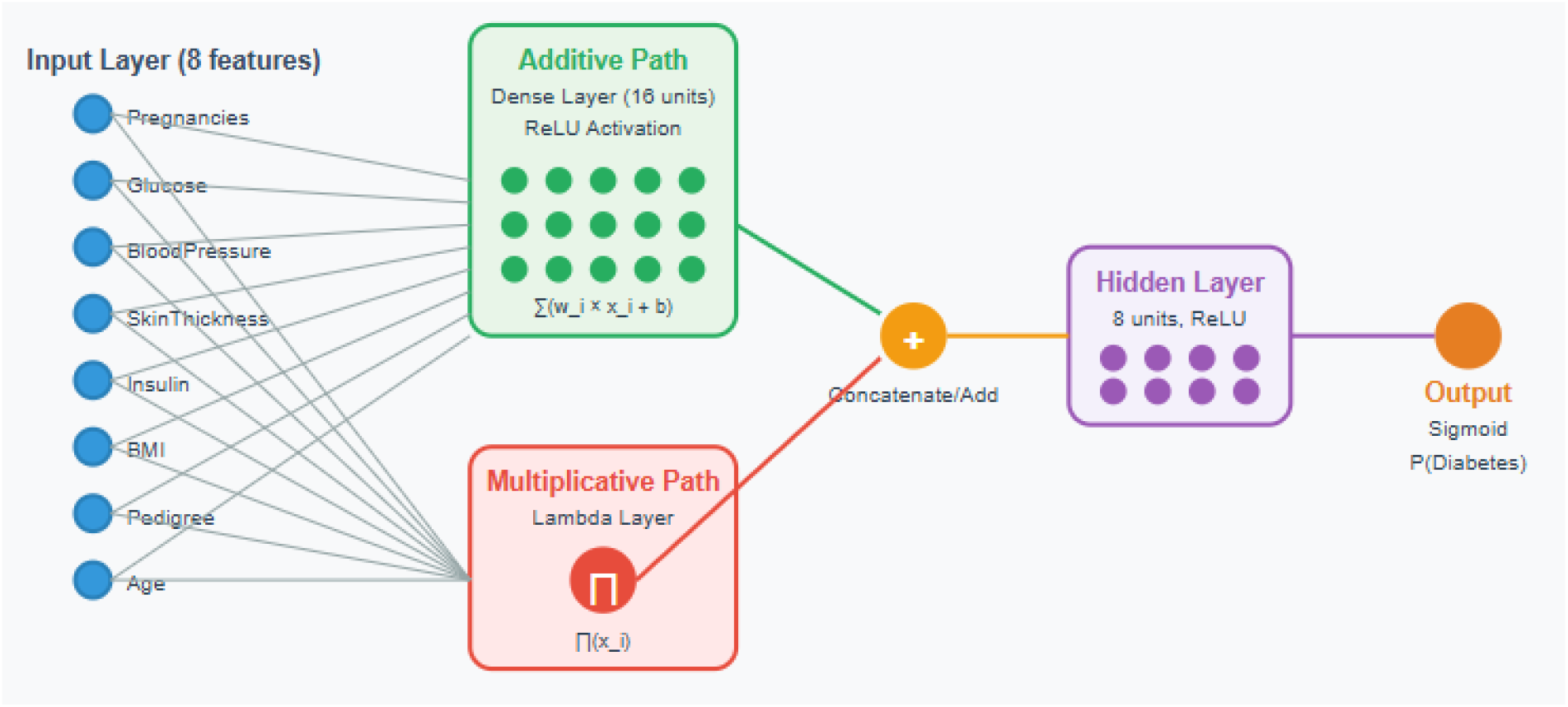
Architectural schematic of the proposed Additive and Multiplicative Neurons Network (AMNN). The architecture integrates two parallel computational pathways: (i) an additive neuron path, which performs a weighted summation of the input signals, and (ii) a multiplicative neuron path, which computes the product of all input features. The outputs of these two streams are subsequently combined and passed through a hidden layer with ReLU activation, followed by a sigmoid-activated output layer for binary classification.

## 3. RESULTS

In this study, the performance of various machine learning algorithms used in diabetes classification was compared. Within the scope of the analysis, 70% of the dataset was allocated for training and 30% for testing, followed by the application of 5-fold cross-validation. Different algorithms such as LR, KNN, MLP, XGB, ET, GNB, KAN and AMNN were evaluated in terms of accuracy, precision, recall, F1-score, and the AUC-ROC. MLP models were tested with different configurations. For instance, MLP-2 used a two hidden layer (100 and 50 neurons), the ReLU activation function, and the “Adam” optimization algorithm. In the KAN model, spline-based activation functions and 10 hidden layer units were employed. In the AMNN model, a hybrid deep learning architecture was constructed that combines additive and multiplicative neurons to better capture both linear and nonlinear relationships. The model consisted of two parallel processing paths: a summation path using ReLU-activated dense neurons (16 units) and a multiplicative path using a custom Lambda layer. These two paths were merged and followed by a hidden layer with 8 neurons (ReLU) and a sigmoid output neuron for binary classificat ion. The AMNN model was compiled using the binary cross-entropy loss function and Adam optimizer, and trained over 100 epochs with a batch size of 32. Performance evaluation was conducted on the test dataset, with performance measures metrics being calculated. Table 5 provides a summary of the comparative performance of all models.

**Table 5.** Performance metrics of selected machine learning algorithms (LR: Logistic Regression, MLP: Multi-Layer Perceptron, XGboost: Extreme Gradient Boosting, ET: Extra Trees, KNN: K-Nearest Neighbors, GNB: Gaussian Naive Bayes, KAN: k-Approximate Nearest, AMNN: Additive and Multiplicative Neurons.)

| Classification Report | Accuracy | Precision | Recall | F1-score | AUC-ROC |
| --- | --- | --- | --- | --- | --- |
| LR | 0.7316 | 0.7386 | 0.7316 | 0.7342 | **0.8347** |
| MLP-2 | 0.7359 | 0.7482 | 0.7359 | 0.7398 | 0.7888 |
| XGboost | 0.7489 | 0.7520 | 0.7489 | 0.7503 | 0.8332 |
| ET | 0.7489 | 0.7463 | 0.7489 | 0.7474 | 0.8286 |
| KNN | 0.7489 | **0.7755** | 0.7489 | 0.7542 | 0.8028 |
| GNB | 0.7273 | 0.7354 | 0.7273 | 0.7302 | 0.8015 |
| KAN | 0.7272 | 0.7185 | 0.7272 | 0.7186 | 0.8225 |
| AMNN | **0.7576** | 0.7733 | **0.7576** | **0.7618** | 0.8206 |

Examining Table 5 reveals that the AMNN model exhibited the best overall performance among the listed models, achieving the highest accuracy (0.7576), recall (0.7576), and F1-score (0.7618). It also delivered a strong precision score (0.7733) and a competitive AUC-ROC value of 0.8206. These results highlight AMNN’s effectiveness in classification tasks, particularly its ability to balance sensitivity and specificity. The model’s high precision suggests strong control over false positives, which is valuable in medical diagnostics. This success is likely due to AMNN’s unique hybrid architecture, combining additive and multiplicative neurons for capturing both linear and nonlinear data relationships.

Following AMNN, the XGBoost, ET, and KNN models formed a mid-performance group, all attaining an accuracy of 0.7489. Among them, KNN stood out with the highest precision (0.7755), indicating reliable positive class identification. XGBoost and ET had competitive F1-scores (0.7503 and 0.7474, respectively), and solid AUC-ROC values above 0.82, reflecting good classification capacity.

The MLP-2 model, with an accuracy of 0.7359 and AUC-ROC of 0.7888, demonstrated moderate performance. LR (Logistic Regression) showed similar trends, achieving an AUC-ROC of 0.8347, the highest among all models, despite its lower accuracy (0.7316), highlighting its strength in distinguishing classes across thresholds.

Models such as GNB, KAN, and LR ranked at the bottom in terms of accuracy and F1-score, though their AUC-ROC values remained above 0.80. KAN, for instance, had a relatively low accuracy (0.7272), but its AUC-ROC (0.8225) suggests stable probability predictions, making it potentially useful in threshold-sensitive applications.

Overall, the results indicate that the AMNN model is a strong alternative to conventional classifiers, demonstrating high accuracy and precision. It is especially suitable for applications where reducing false positives is of primary importance. Ensemble-based models and tree-based learners (like XGBoost and ET) also proved effective, while simpler models like LR and GNB, despite lower classification metrics, still offer value in terms of interpretability and AUC-based ranking.

## 4. DISCUSSION

This research aimed to assess the diagnostic capabilities of several machine learning algorithms for diabetes, with the aim of determining which model proved most effective. Using the PIMA Indians Diabetes dataset, the analysis revealed that the AMNN models achieved the highest performance. These models demonstrated the highest accuracy and AUC-ROC scores, indicating that deep learning and ensemble-based methods are particularly effective for diabetes classification.

The application of XAI methods such as SHAP and LIME provided valuable insights into model decision-making processes. These methods highlighted that glucose level, BMI, and age are the most critical features in predicting diabetes. Among these, glucose level was found to be the strongest predictor, significantly influencing the model’s ability to classify individuals as diabetic or non-diabetic.

This finding aligns with clinical evidence, as elevated blood glucose levels are a primary diagnostic criterion for diabetes and play a central role in disease progression [35]. BMI, another key factor, is strongly associated with insulin resistance and metabolic dysfunction, making it a crucial indicator for early intervention [36]. Age-related changes in glucose metabolism and pancreatic function further emphasize the importance of age as a predictive variable in diabetes risk assessment [37]. Other important features included Diabetes Pedigree Function and the number of pregnancies. A strong family history of diabetes, as indicated by the Diabetes Pedigree Function, suggests a genetic predisposition, which is critical for identifying at-risk individuals before clinical symptoms appear [38]. Additionally, the number of pregnancies may reflect the impact of gestational diabetes, a known risk factor for type 2 diabetes in later life [39]. Recognizing these risk factors through machine learning models allows for earlier lifestyle modifications, targeted screening programs, and personalized treatment plans [40,41]. However, features like blood pressure, skin thickness, and insulin levels had a lower impact on the model’s predictions. While these factors may not be the strongest predictors, they still contribute to the overall assessment of metabolic health and should not be overlooked in clinical practice [42].

These findings are largely consistent with previous studies in the literature. For instance, Kayaer and Yıldırım [4] achieved 80.21% accuracy using a GRNN on the same dataset, while Patra and Khuntia [9] improved the KNN classifier to achieve 83.2% accuracy. Similarly, Mousa et al. [10] found that LSTM achieved the highest accuracy (85%) among various models applied to the PIMA dataset. In this study, although the LSTM model was not implemented, a number of classical and hybrid machine learning models were tested, including ensemble and deep learning-based approaches.

According to the experimental results, the best performance was achieved by the AMNN model, with an accuracy of 75.76%, a recall of 75.76%, and an F1-score of 76.18%. These metrics demonstrate a competitive performance compared to several previously reported models. Furthermore, ensemble models such as XGBoost and ET (Extra Trees) also showed promising results, with accuracy values of 74.89%, aligning with the findings of Yangın [6] and Chang et al. [11], who emphasized the strength of ensemble-based methods like XGBoost and RF. Additionally, KNN achieved a solid precision of 77.55%, which is consistent with the enhanced version presented by Patra and Khuntia [9].

On the other hand, logistic regression (LR) yielded a relatively lower accuracy of 73.16% but reached the highest AUC-ROC value of 0.8347, indicating a good balance between sensitivity and specificity. This result also resonates with the findings by Chang et al. [11], where interpretable models like Naïve Bayes and decision trees were found to be effective with fewer features.

In summary, the findings of this study confirm the effectiveness of neural and ensemble learning approaches for diabetes classification and reinforce the idea that hybrid models like AMNN can provide robust performance across multiple evaluation metrics. The consistency of these results with prior research supports the continued use and development of such models in medical diagnostic tasks involving structured datasets like PIMA.

Although the results are encouraging, there are some limitations to this study.The PIMA Indians Diabetes dataset contains only 768 observations, which limits the generalizability of the findings. Moreover, the dataset is imbalanced, containing fewer diabetic cases relative to non-diabetic ones. To mitigate this issue, the SMOTE oversampling technique was employed. Nevertheless, it remains important to evaluate how such oversampling approaches affect the model’s ability to generalize to real-world scenarios. Future research could benefit from larger and more balanced datasets to enhance model reliability.

Another limitation is that the PIMA dataset includes only female participants, which prevents an analysis of gender-based differences in diabetes prediction. Future studies could incorporate datasets that include both male and female individuals to examine potential gender-based variations in diabetes risk factors. Moreover, this study incorporated both conventional machine learning models and deep learning algorithms, involving a novel hybrid approach (AMNN). For future research, exploring more advanced hybrid models, transfer learning strategies, or federated learning techniques could further enhance diabetes classification performance. Additionally, implementing these models in real-world clinical decision support systems could help validate their practical applicability.

## 5. CONCLUSION

This study aims to improve diabetes classification using various machine learning models, while also identifying the most influential variables through explainable AI (XAI) and feature selection methods. A key innovation of the study is the implementation of the AMNN model, a novel artificial neural network (ANN) architecture that outperformed all other models in diabetes prediction. In addition, the study explores the KAN model, another recent ANN approach, which offers a distinctive design through learnable edge-based activation functions.

The experimental results demonstrate that the AMNN model achieved the highest classification performance, with an accuracy of 75.76%, recall of 75.76%, and F1-score of 76.18%, outperforming other models in overall predictive capability. Although LR (Logistic Regression) recorded the highest AUC-ROC value (0.8347), indicating strong discriminatory power, the overall balanced performance of AMNN across all metrics makes it the most effective model in this study.

Ensemble models like XGBoost and ET (Extra Trees) also showed competitive results, confirming their robustness in medical classification tasks. While the KAN model did not surpass AMNN, it achieved a respectable accuracy of 72.72% and AUC-ROC of 0.8225. Its innovative use of learnable activation functions at the edges instead of traditional fixed-node activations offers greater adaptability and interpretability, marking it as a promising direction for future studies.

In terms of feature importance, both feature selection and XAI techniques consistently highlighted glucose level, BMI, and age as the most significant predictors of diabetes. Additionally, Diabetes Pedigree Function and number of pregnancies were found to contribute meaningfully to classification outcomes, which aligns with established medical knowledge.

Despite these encouraging findings, some limitations remain. The dataset is relatively small, includes only female participants of Pima Indian heritage, and is class-imbalanced, which may limit generalizability. Future research should aim to address these limitations by utilizing larger, more diverse datasets and integrating advanced techniques such as hybrid learning, transfer learning, or federated learning to improve model accuracy and robustness.

In conclusion, the results confirm that machine learning, particularly deep learning and ensemble-based methods like AMNN and XGBoost, hold great potential for early diabetes diagnosis. The integration of XAI techniques enhances model transparency, facilitating trust and interpretability in clinical decision-making. Moreover, innovative models like KAN warrant further exploration, especially in hybrid configurations, to advance diabetes prediction and support personalized healthcare applications.

## Data Availability

All data analyzed in this study were obtained from the publicly available PIMA Indians Diabetes dataset (https://www.kaggle.com/datasets/uciml/pima-indians-diabetes-database
). No additional data were generated

